# Association of Cancer with Risk and Mortality of COVID-19: Results from the UK Biobank

**DOI:** 10.1101/2020.07.10.20151076

**Authors:** Zhuqing Shi, W. Kyle Resurreccion, Chi-Hsiung Wang, Jun Wei, Rong Na, S. Lilly Zheng, Liana K. Billings, Brian T. Helfand, Janardan Khandekar, Jianfeng Xu

## Abstract

Although cancer has been associated with COVID-19 risk and mortality in hospital-based studies, few population-based studies have been reported. Utilizing data from the UK Biobank (UKB), a population-based prospective cohort, we formally tested the association of over 44 different types of cancer with COVID-19 infection and mortality among 7,661 subjects who were tested by June 17, 2020. Compared to non-cancer subjects, cancer subjects (N=1,521) had significantly lower overall risk for COVID-19 infection [odds ratio (OR) and 95% confidence interval (CI): 0.79 (0.68-0.92), *P*=2.60E-03]. However, a trend of higher risk for COVID-19 mortality was found among 256 COVID-19 positive cancer patients, especially for hematologic cancers such as non-Hodgkin lymphoma [3.82 (1.17-12.01), P=0.02]. In cancer patients, while few demographic, lifestyle, genetic and comorbidity factors predicted risk for COVID-19 infection, older age, male sex, heart disease and hypertension significantly predicted COVID-19 mortality. The lower risk for COVID-19 infection is likely due to extra caution in COVID-19 prevention and more testing among cancer patients, an encouraging finding that demonstrates the feasibility of intervention. These results, if confirmed in future releases of UKB data and other independent populations, may provide guidance for COVID-19 prevention and treatment among cancer patients.

## Introduction

The coronavirus disease 2019 (COVID-19), caused by severe acute respiratory syndrome coronavirus 2 (SARS-CoV-2), is a global pandemic. Based on the Johns Hopkins Coronavirus Resource Center on July 7, 2020, 11,662,574 confirmed cases of COVID-19 worldwide were reported.[1] In the United Kingdom (UK), 287,291 cases and 44,321 deaths from COVID-19 were documented.

A number of risk factors and pre-existing comorbid conditions such as cardiovascular disease, diabetes, chronic respiratory disease, hypertension and cancer have been associated with risk for COVID-19 infection and severity.[2-4] Specifically, several studies on the characteristics of COVID-19 among cancer patients were reported. In a prospective observational study from 55 cancer centers in the UK, COVID-19 mortality in cancer patients was associated with age, gender, and comorbidities.[5] A study based on the COVID-19 and Cancer Consortium (CCC19) from the United States, Canada and Spain reported a high mortality rate among patients with both cancer and COVID-19, with mortality being associated with general and cancer-specific risk factors.[6] A higher percentage of severe COVID-19 patients, including those hospitalized, who required mechanical ventilation and died within 30 days, was reported in a study at Memorial Sloan Kettering Cancer Center.[7] However, to date, these reported studies on cancer and COVID-19 are all hospital-based. As such, it is yet unknown whether cancer patients in the general population have higher risk and mortality for COVID-19 compared to individuals without cancer.

The goal of this study was to compare COVID-19 risk and mortality between subjects with and without a cancer diagnosis in the UK Biobank (UKB), a large population-based cohort in the UK. Furthermore, leveraging the comprehensive data on demographics, genetics, lifestyle, and disease phenotypes in the UKB, we performed additional analyses to compare the impact of candidate risk factors and comorbid conditions on COVID-19 positivity and mortality in subjects with and without cancer.

## Methods

The UKB is a population-based study biorepository for approximately 500,000 individuals aged 40-69 at recruitment in 2006-2010 from across the UK.[8] Extensive phenotypic and health-related information is available for each participant, including questionnaire, biological measurements, lifestyle indicators, and biomarkers in blood and urine. Follow-up information is provided by linking health and medical records. Cancer status, related comorbidities (chronic respiratory diseases, obesity, diabetes, hypertension, stroke), and risk factors can be obtained through the International Classification of Diseases-10 (ICD-10) codes, cancer registry, death registry, and self-report. More than 44 types of cancer were recorded in UKB.

COVID-19 test results for the UKB participants are provided by Public Health England (for participants resident in England). This information is updated weekly. Additional COVID-19 data is also collected through a) general practitioner (primary care) data, b) hospital inpatient data, c) death data, and d) critical care data (for COVID-19 positive patients), which are updated monthly.

UK Biobank Axiom SNP array data is available for all subjects. Blood type (ABO) were inferred from the genotypes of rs8176746 and rs8176719, and APOE alleles were inferred from the genotype of rs7412 and rs429358 (ε2=T/T, ε3=C/T and ε4=C/C).[9]

Association tests were performed using univariate and multivariable (adjusting for age and gender) logistic regression. Differences in variables between groups were tested using t-test (for continuous variables) and Chi-square test (for categorical variables) or the Fisher’ s exact test when the numbers are small (expected frequencies <5). Statistical analyses were performed by R v3.5.2, and two-tailed *P*<0.05 was considered statistically significant.

## Results

By June 17, 2020, COVID-19 test results were available for 7,661 subjects, approximately 1.5% of UKB subjects. Among these subjects, 1,521 had a diagnosis for one or more of 44 different types of cancer. Subjects with cancer had significantly older mean age (61.58) than non-cancer subjects (56.99), *P*<0.0001. Cancer patients also had a significantly higher proportion of males (54.11%) than non-cancer subjects (47.92%), *P*<0.0001.

Among all subjects, 1,562 (20.39%) tested positive for COVID-19, of which, 206 (13.19%) died (**Table 1**). The COVID-19 positive rate was lower among cancer patients (16.83%) than non-cancer subjects (21.27%). Compared to non-cancer subjects, cancer patients were significantly associated with lower positive rates for COVID-19 in both univariate analysis [OR= 0.75 (0.65-0.87), P=1.24E-04] and multivariable analysis adjusting for age and gender [OR=0.79 (0.68-0.92), *P*=2.60E-03]. For mortality among subjects positive for COVID-19, however, the rate was higher in cancer patients (17.97%) than non-cancer subjects (12.25%). Compared to non-cancer subjects, cancer patients were significantly associated with higher COVID-19 mortality in the univariate analysis [OR=1.57 (1.09-2.23), P=0.01], but the association diminished after adjusting for age and gender [OR=1.04 (0.71-1.51), *P*=0.83].

**Table 1.**
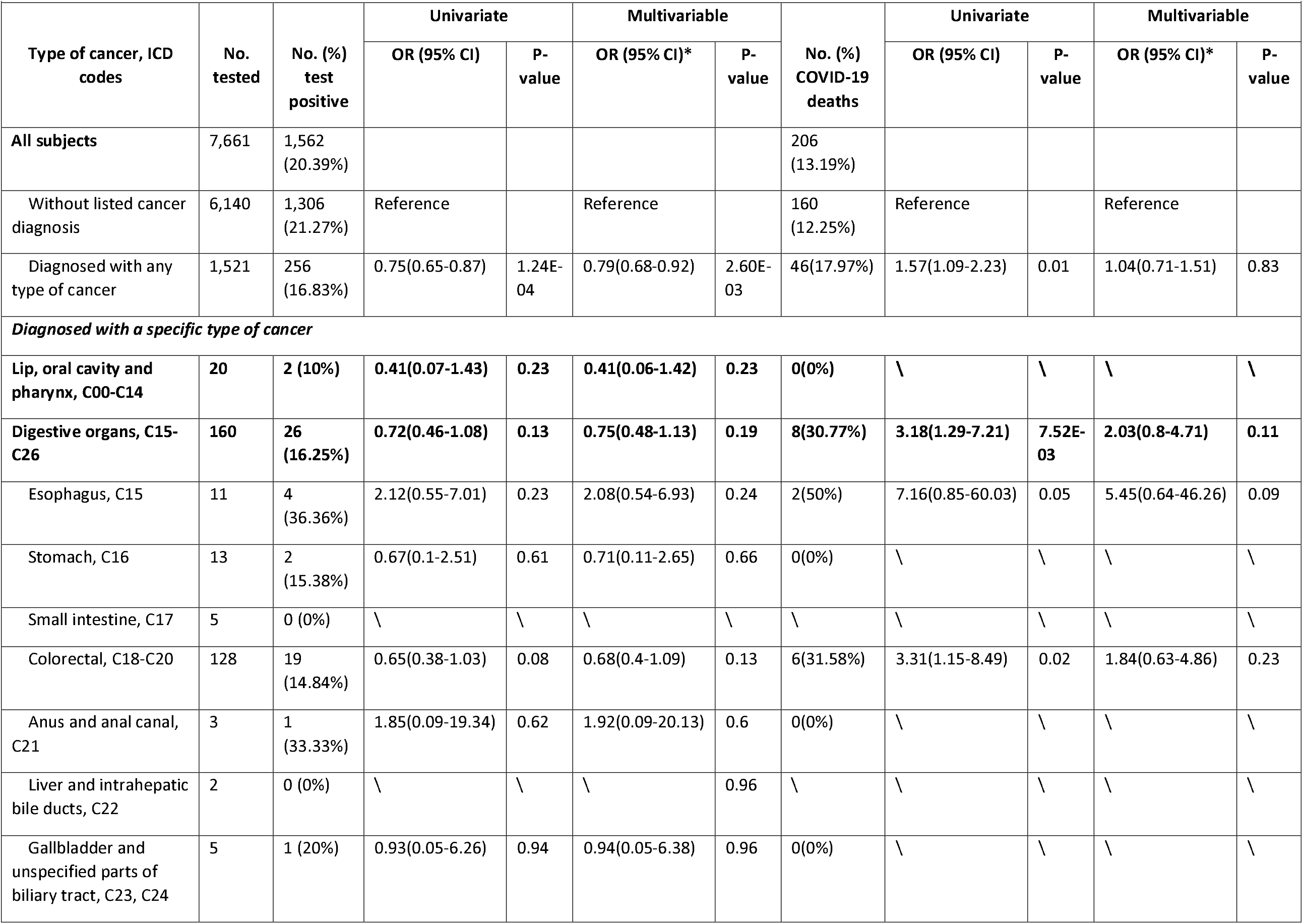
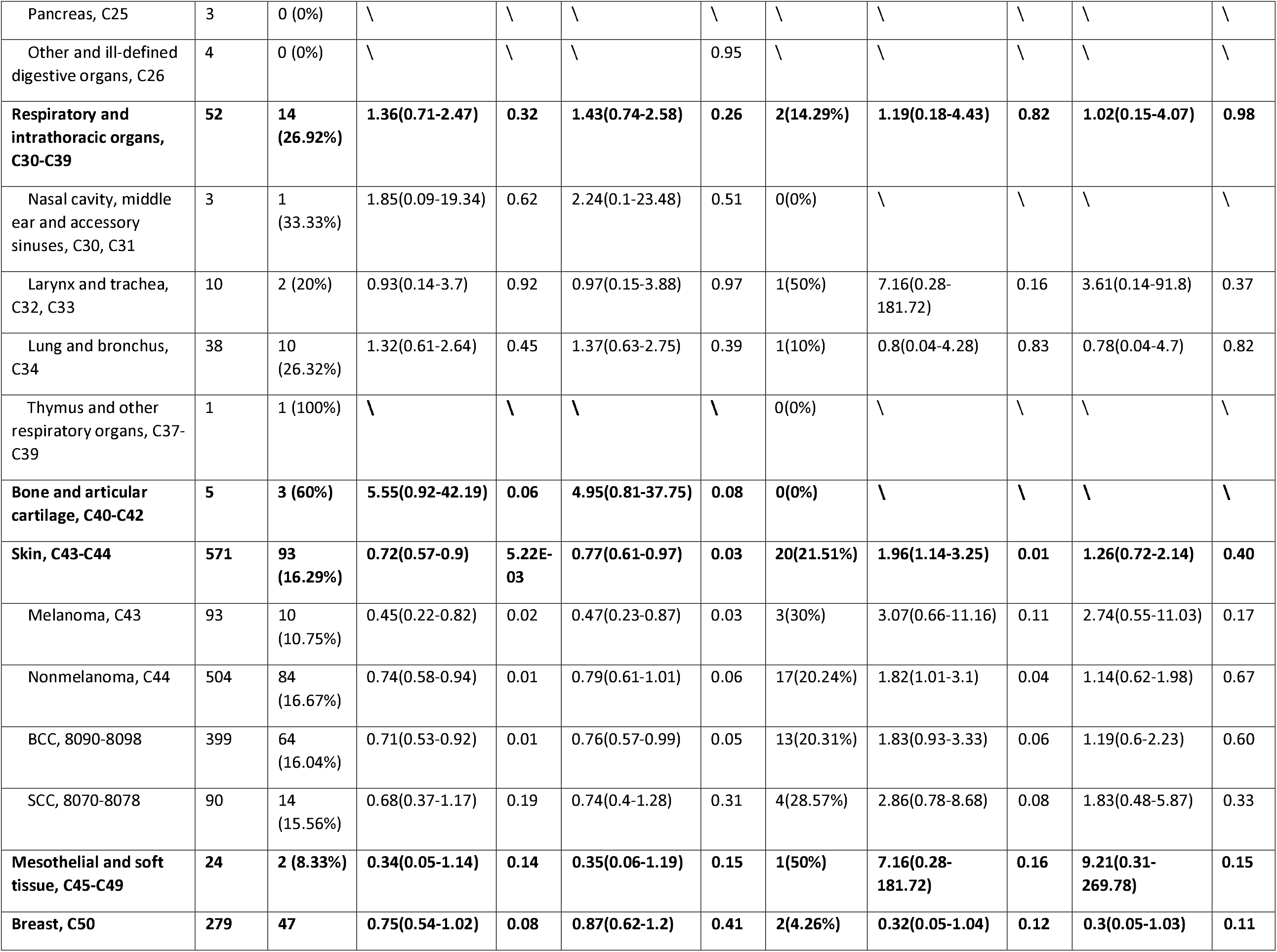
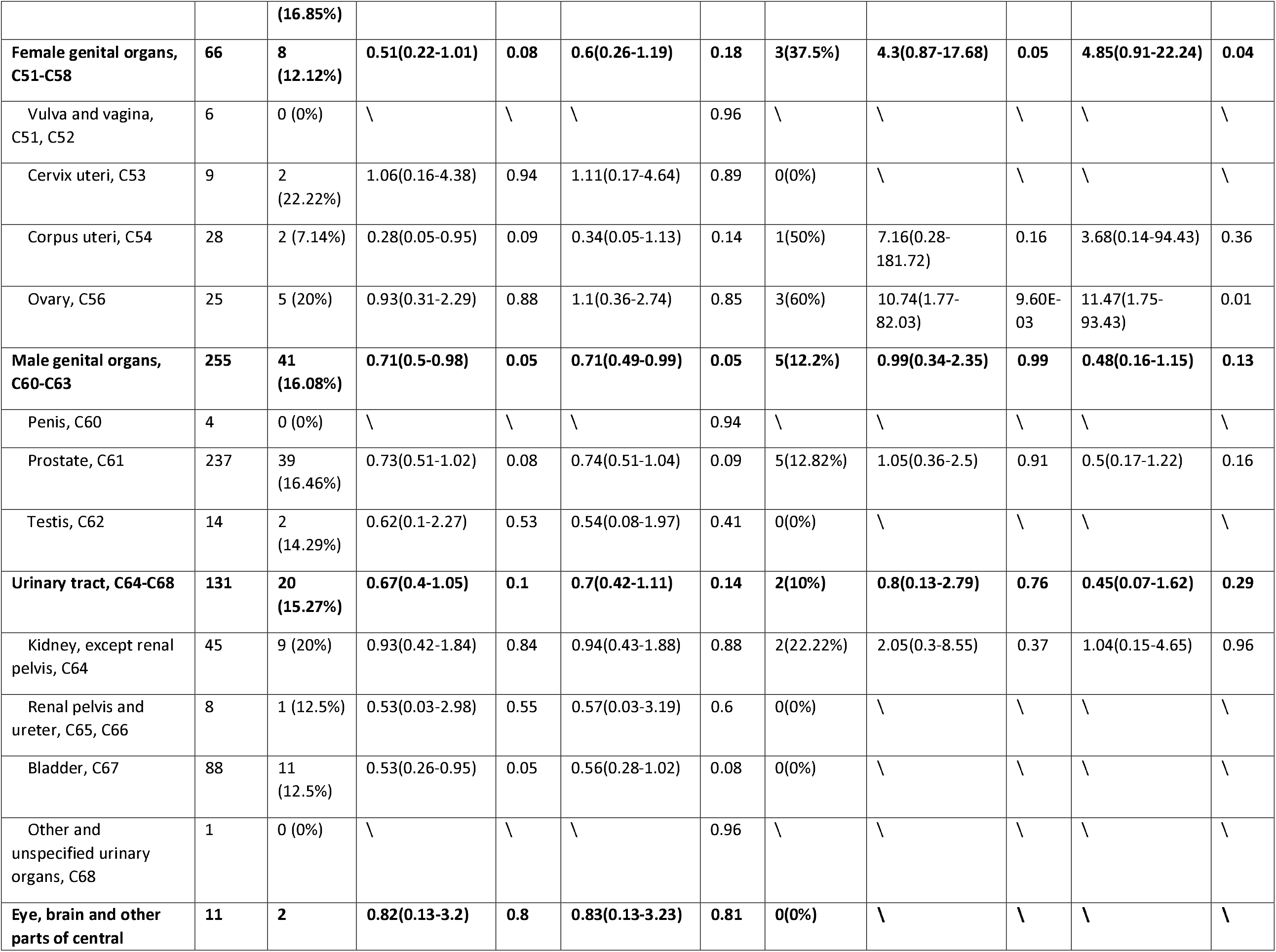
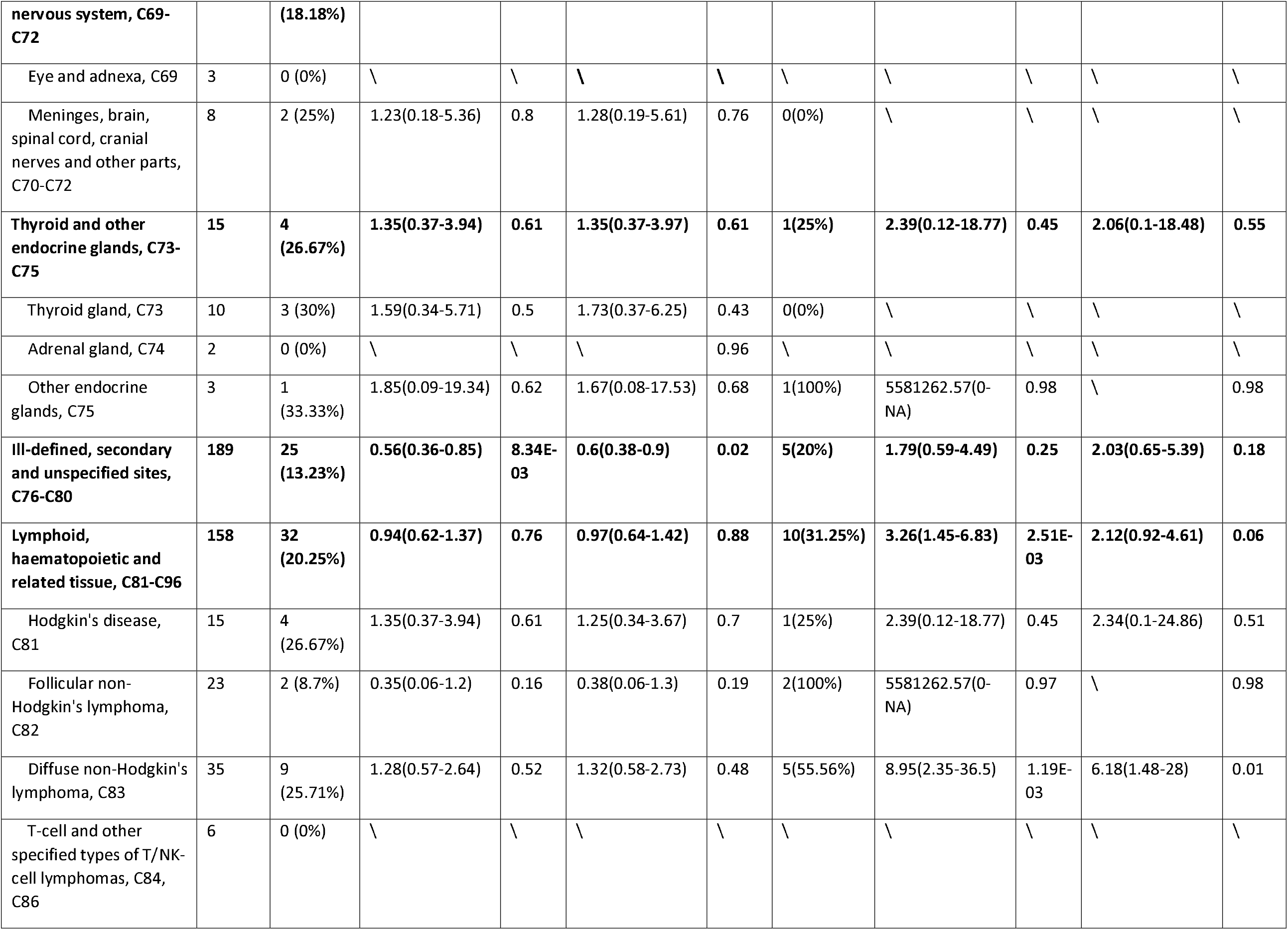
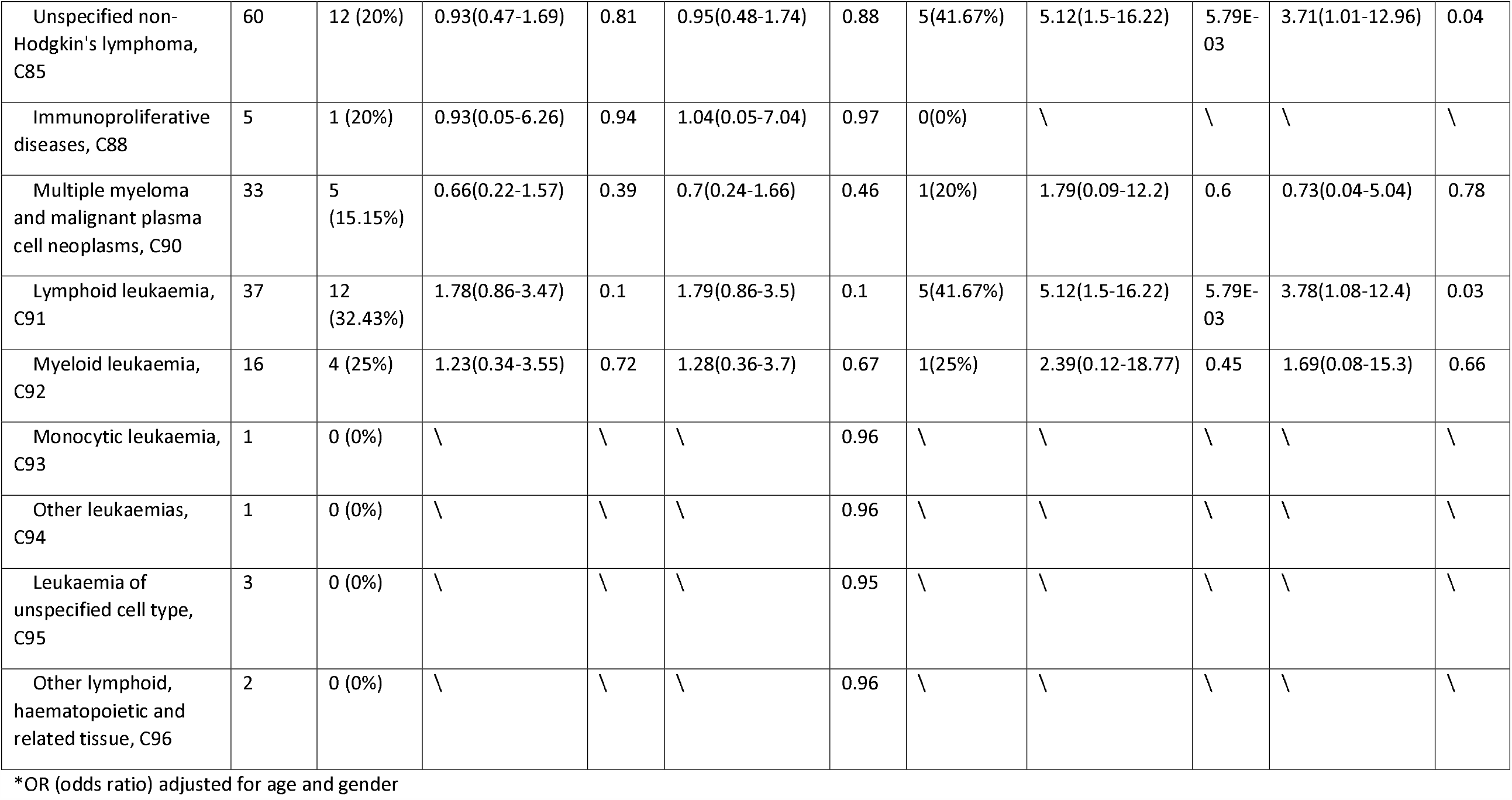
Association of cancer diagnosis with COVID-19 infection and mortality in UK Biobank

When examining specific types of cancer, a lower COVID-19 positive rate was found for the vast majority of cancers in various organ systems including digestive, skin, breast, female genital, male genital, urinary track, brain/central nervous system, and hematological tissues. However, higher COVID-19 positive rates were found among those with cancer in the respiratory/intrathoracic organs, bone/articular cartilage, and thyroid and other endocrine glands, although the differences were not statistically significant.

Among COVID-19 positive patients, significantly higher COVID-19 mortality was found for patients with many types of cancer in the univariate analyses. Most of these associations attenuated in multivariable analysis (**Table 1**). However, significantly higher COVID-19 mortality was found for several types of hematologic cancers in multivariable analyses adjusting for age and gender. For example, for non-Hodgkin’ s lymphoma (ICD-10 codes C82, C83, C85) patients, the odds for COVID-19 mortality was significantly higher [OR=3.82 (1.17-12.01), P=0.02] than non-cancer patients.

Finally, we tested the association between candidate predictors (demographic, lifestyle, genetic and comorbidity) with COVID-19 positivity and mortality separately in cancer and non-cancer patients (**Table 2**). Several observations were noted. First, among cancer patients, few predictors except for minority race were significantly associated with a higher positivity rate. In contrast, among the cancer patients who were positive for COVID-19, multiple predictors including older age, male sex, and comorbidities such as heart diseases and hypertension were significantly associated with COVID-19 death. Higher mortality risk was also associated with two other comorbidities (obesity and diabetes), although not statistically significant. Second, candidate genetic predictors (ABO blood type and APOE) were not significantly associated with COVID-19 positivity and mortality, although a modest association between the homozygous APOE ε4 allele and COVID-19 positivity/mortality was found. Third, the above two observations were not unique to cancer patients and were also found in non-cancer patients, although there were more statistically significant associations due to larger sample size. For example, multiple comorbidities were associated with COVID-19 mortality but not positivity.

**Table 2.** Characteristics of cancer patients in the UK Biobank

| **Table 2.** Characteristics of cancer patients in the UK Biobank |  |  |  |  |  |  |  |  |  |  |  |  |  |  |  |  |
| --- | --- | --- | --- | --- | --- | --- | --- | --- | --- | --- | --- | --- | --- | --- | --- | --- |
|  | Cancer patients, N=1,521 |  |  |  | Cancer patients, COVID-19 positive, N=256 |  |  |  | Non-cancer subjects, N=6,140 |  |  |  | Non-cancer subjects, COVID-19 positive, N=1,306 |  |  |  |
|  | Test negative | Test positive | OR (95% CI) | P-value | Alive | Died of COVID-19 | OR (95% CI) | P-value | Test negative | Test positive | OR (95% CI) | P-value | Alive | Died of COVID-19 | OR (95% CI) | P-value |
| **No. (%) of subjects** | 1,265 (83.17%) | 256 (16.83%) |  |  | 210 (82.03%) | 46 (17.97%) |  |  | 4,834 (78.73%) | 1,306 (21.27%) |  |  | 1,146 (87.75%) | 160 (12.25%) |  |  |
| **Demographic variables** |  |  |  |  |  |  |  |  |  |  |  |  |  |  |  |  |
| **Age, mean (IQR), year** | 61.62 (58.5-66.5) | 61.36 (56.5-67.5) | 0.99 (0.98-1.01) | 0.58 | 60.47 (55.5-66.5) | 65.41 (63.5-68.5) | 1.15 (1.08-1.22) | 4.87E-10 | 57.23 (49.5-64.5) | 56.11 (47.5-64.5) | 0.99 (0.98-0.99) | 7.72E-05 | 55.24 (46.5-63.5) | 62.33 (58.5-67.5) | 1.11 (1.08-1.13) | 2.04E-28 |
| **Gender, No. (%) male** | 673 (53.2%) | 150 (58.59%) | 1.24 (0.95-1.63) | 0.13 | 114 (54.29%) | 36 (78.26%) | 3.03 (1.43-6.43) | 4.73E-03 | 2,276 (47.08%) | 666 (51%) | 1.17 (1.03-1.32) | 0.01 | 560 (48.87%) | 106 (66.25%) | 2.05 (1.45-2.91) | 5.43E-05 |
| **Race** |  |  |  | 3.59E-03 |  |  |  | 1 |  |  |  | 1.61E-11 |  |  |  | 6.36E-03 |
| White, No. (%) | 1223 (96.91%) | 234 (92.49%) | Ref |  | 192 (92.31%) | 42 (93.33%) | Ref |  | 4434 (92.34%) | 1119 (86.28%) | Ref |  | 982 (86.29%) | 137 (86.16%) | Ref |  |
| Black, No. (%) | 17 (1.35%) | 10 (3.95%) | 3.07 (1.54-5.91) |  | 8 (3.85%) | 2 (4.44%) | 1.14 (0.24-3.84) |  | 122 (2.54%) | 73 (5.63%) | 2.37 (1.84-3.04) |  | 57 (5.01%) | 16 (10.06%) | 2.01 (1.21-3.23) |  |
| Asian, No. (%) | 10 (0.79%) | 6 (2.37%) | 3.14 (1.27-7.27) |  | 5 (2.4%) | 1 (2.22%) | 0.91 (0.09-4.46) |  | 135 (2.81%) | 67 (5.17%) | 1.97 (1.52-2.52) |  | 61 (5.36%) | 6 (3.77%) | 0.71 (0.32-1.37) |  |
| Other, No. (%) | 12 (0.95%) | 3 (1.19%) | 1.31 (0.39-3.49) |  | 3 (1.44%) | 0 (0%) | \ |  | 111 (2.31%) | 38 (2.93%) | 1.36 (0.98-1.84) |  | 38 (3.34%) | 0 (0%) | 0 (0-0.58) |  |
| **Lifestyle risk factors** |  |  |  |  |  |  |  |  |  |  |  |  |  |  |  |  |
| **BMI, mean (IQR)** | 28.13 (24.79-30.67) | 28.71 (25.15-31.03) | 1.02 (1-1.04) | 0.09 | 28.47 (24.92-30.94) | 29.84 (25.64-32.77) | 1.05 (1-1.1) | 0.11 | 28.25 (24.62-31.02) | 28.94 (25.18-31.8) | 1.02 (1.01-1.03) | 3.71E-05 | 28.75 (25.03-31.47) | 30.29 (26.14-33.32) | 1.05 (1.02-1.07) | 1.01E-03 |
| **Smoking, No. (%) previous/current** | 681 (54.13%) | 154 (60.87%) | 1.32 (1-1.74) | 0.06 | 128 (61.54%) | 26 (57.78%) | 0.86 (0.44-1.65) | 0.76 | 2,463 (51.28%) | 635 (49.15%) | 0.92 (0.81-1.04) | 0.18 | 535 (47.22%) | 100 (62.89%) | 1.89 (1.35-2.67) | 2.98E-04 |
| **Genetic risk factors** |  |  |  |  |  |  |  |  |  |  |  |  |  |  |  |  |
| **ABO blood type, No. (%)** |  |  |  | 0.84 |  |  |  | 0.19 |  |  |  | 0.72 |  |  |  | 0.71 |
| AB/B | 161 (13.44%) | 29 (12.03%) | Ref |  | 20 (10.2%) | 9 (20%) | Ref |  | 657 (14.26%) | 189(15.17%) | Ref |  | 166 (15.19%) | 23 (15.03%) | Ref |  |
| A | 529 (44.16%) | 108 (44.81%) | 1.13 (0.79-1.67) |  | 89 (45.41%) | 19 (42.22%) | 0.47 (0.22-1.06) |  | 2069 (44.91%) | 556 (44.62%) | 0.93 (0.8-1.09) |  | 492 (45.01%) | 64 (41.83%) | 0.94 (0.62-1.46) |  |
| O | 508 (42.4%) | 104 (43.15%) | 1.14 (0.79-1.67) |  | 87 (44.39%) | 17 (37.78%) | 0.43 (0.2-0.98) |  | 1881 (40.83%) | 501 (40.21%) | 0.93 (0.79-1.09) |  | 435 (39.8%) | 66 (43.14%) | 1.1 (0.72-1.7) |  |
| **APOE, No. (%)** |  |  |  | 0.22 |  |  |  | 0.12 |  |  |  | 2.03E-03 |  |  |  | 0.1 |
| 0 ε4 | 753<br>(74.48%) | 137<br>(69.54%) | Ref |  | 108<br>(67.92%) | 29<br>(76.32%) | Ref |  | 2837<br>(72.46%) | 739<br>(69.26%) | Ref |  | 658<br>(70.3%) | 81<br>(61.83%) | Ref |  |
| 1 ε4 | 234<br>(23.15%) | 52<br>(26.4%) | 1.22<br>(0.91-1.63) |  | 46<br>(28.93%) | 6<br>(15.79%) | 0.49<br>(0.21-1.03) |  | 978<br>(24.98%) | 280<br>(26.24%) | 1.1<br>(0.96-1.25) |  | 239<br>(25.53%) | 41<br>(31.3%) | 1.39<br>(0.99-1.95) |  |
| 2 ε4 | 24<br>(2.37%) | 8<br>(4.06%) | 1.83<br>(0.88-3.54) |  | 5<br>(3.14%) | 3 (7.89%) | 2.23<br>(0.58-7.62) |  | 100<br>(2.55%) | 48<br>(4.5%) | 1.84<br>(1.36-2.47) |  | 39<br>(4.17%) | 9 (6.87%) | 1.87<br>(0.95-3.44) |  |
| **Comorbidity** |  |  |  |  |  |  |  |  |  |  |  |  |  |  |  |  |
| **Chronic obstructive pulmonary disease, No. (%)** | 265<br>(20.95%) | 51<br>(19.92%) | 0.94<br>(0.67-1.31) | 0.78 | 41<br>(19.52%) | 10<br>(21.74%) | 1.14<br>(0.53-2.5) | 0.89 | 778<br>(16.09%) | 188<br>(14.4%) | 0.88<br>(0.74-1.04) | 0.15 | 154<br>(13.44%) | 34<br>(21.25%) | 1.74<br>(1.15-2.63) | 0.01 |
| **Asthma, No. (%)** | 237<br>(18.74%) | 36<br>(14.06%) | 0.71<br>(0.49-1.04) | 0.09 | 32<br>(15.24%) | 4 (8.7%) | 0.53<br>(0.18-1.58) | 0.35 | 811<br>(16.78%) | 204<br>(15.62%) | 0.92<br>(0.78-1.09) | 0.34 | 183<br>(15.97%) | 21<br>(13.12%) | 0.8<br>(0.49-1.29) | 0.42 |
| **Heart disease, No. (%)** | 719<br>(56.84%) | 140<br>(54.69%) | 0.92<br>(0.7-1.2) | 0.57 | 107<br>(50.95%) | 33<br>(71.74%) | 2.44<br>(1.22-4.9) | 0.02 | 1,950<br>(40.34%) | 532<br>(40.74%) | 1.02<br>(0.9-1.15) | 0.82 | 430<br>(37.52%) | 102<br>(63.75%) | 2.93<br>(2.08-4.13) | 4.40E-10 |
| **Stroke, No. (%)** | 59<br>(4.66%) | 11<br>(4.3%) | 0.92<br>(0.48-1.77) | 0.93 | 9<br>(4.29%) | 2 (4.35%) | 1.02<br>(0.21-4.86) | 1 | 188<br>(3.89%) | 53<br>(4.06%) | 1.05<br>(0.77-1.43) | 0.84 | 44<br>(3.84%) | 9 (5.62%) | 1.49<br>(0.71-3.12) | 0.39 |
| **Hypertension, No. (%)** | 648<br>(51.23%) | 137<br>(53.52%) | 1.1<br>(0.84-1.43) | 0.55 | 106<br>(50.48%) | 31<br>(67.39%) | 2.03<br>(1.03-3.98) | 0.05 | 2,062<br>(42.66%) | 552<br>(42.27%) | 0.98<br>(0.87-1.11) | 0.82 | 449<br>(39.18%) | 103<br>(64.38%) | 2.81<br>(1.99-3.96) | 2.55E-09 |
| **Obesity, No. (%)** | 108<br>(8.54%) | 22<br>(8.59%) | 1.01<br>(0.62-1.63) | 1 | 17<br>(8.1%) | 5<br>(10.87%) | 1.38<br>(0.48-3.97) | 0.75 | 373<br>(7.72%) | 102<br>(7.81%) | 1.01<br>(0.81-1.27) | 0.96 | 80<br>(6.98%) | 22<br>(13.75%) | 2.12<br>(1.28-3.52) | 4.63E-03 |
| **Diabetes, No. (%)** | 183<br>(14.47%) | 44<br>(17.19%) | 1.23<br>(0.86-1.76) | 0.31 | 34<br>(16.19%) | 10<br>(21.74%) | 1.44<br>(0.65-3.17) | 0.49 | 582<br>(12.04%) | 188<br>(14.4%) | 1.23<br>(1.03-1.47) | 0.03 | 145<br>(12.65%) | 43<br>(26.88%) | 2.54<br>(1.72-3.75) | 2.86E-06 |
| **Any 1 comorbidity, No. (%)** | 900<br>(71.15%) | 180<br>(70.31%) | 0.96<br>(0.72-1.29) | 0.85 | 143<br>(68.1%) | 37<br>(80.43%) | 1.93<br>(0.88-4.22) | 0.14 | 2,868<br>(59.33%) | 756<br>(57.89%) | 0.94<br>(0.83-1.07) | 0.36 | 633<br>(55.24%) | 123<br>(76.88%) | 2.69<br>(1.83-3.96) | 3.26E-07 |
| **Two or more comorbidities, No. (%)** | 715<br>(56.52%) | 141<br>(55.08%) | 0.95<br>(0.73-1.23) | 0.91 | 111<br>(52.86%) | 30<br>(65.22%) | 2.01<br>(1.05-4.09) | 0.23 | 2080<br>(43.03%) | 563<br>(43.11%) | 0.97<br>(0.87-1.08) | 0.36 | 461<br>(40.23%) | 102<br>(63.75%) | 3.07<br>(2.21-4.31) | 4.20E-08 |

## Discussion

Several important findings were obtained in this first population-based and observational study that formally assessed the association of cancer with COVID-19 infection and mortality. The first finding that cancer patients had an overall significantly lower infection rate of COVID-19 than non-cancer subjects is novel. It appears contrary to the hypothesis of systemic immunosuppressive state in cancer patients caused by the malignancy and anticancer treatments.[10] However, the finding is not unexpected for this type of observational study due to a combination of several factors: 1) most non-cancer subjects who underwent testing likely had COVID-19 symptoms early in the pandemic when testing was limited; 2) on the other hand, more cancer patients who opted for testing due to extra caution were asymptomatic; and 3) cancer patients practiced stronger personal protection against COVID-19. Nevertheless, this finding is encouraging because it empirically demonstrated that the risk for COVID-19 among cancer patients, even though they may be more susceptible to the disease biologically, can be practically mitigated.

A trend of increased COVID-19 positive rates in patients of several specific types of cancer in three organ systems (respiratory/intrathoracic organs, bone articular cartilage, and thyroid/other endocrine glands) is another novel finding. These results are intriguing considering the overall lower infection rate among cancer patients observed in this cohort. It is noted that these associations did not reach statistical significance. Confirmation in additional studies is needed.

The third major finding that cancer patients have a trend of increased COVID-19 mortality is consistent with previously published studies.[2-7] The confirmation from this population-based prospective study that included a broad range of over 44 cancer types further strengthens the available evidence and makes it more generalizable to cancer patients in the general population. Among specific types of cancer, the higher COVID-19 mortality in patients of several hematologic cancers, especially Non-Hodgkin lymphoma, is noticeable. A similar finding was also reported in the cancer cohort from Memorial Sloan Kettering Cancer Center.[7] The exact mechanism is unknown, but low lymphocyte counts in these patients is suspected. A previous study suggests that COVID-19 patients with more severe symptoms have lower lymphocyte counts caused by the functional exhaustion and apoptosis of cytotoxic T lymphocytes.[11] Another study associated lymphopenia with poorer overall survival for individuals with Non-Hodgkin lymphoma.[12]

The last finding was that multiple demographic and comorbid conditions predicted COVID-19 mortality, but not COVID-19 positivity. The lack of association with COVID-19 positivity is unlikely explained by sample size, because many more subjects were available for testing positivity than mortality. Furthermore, it is interesting to note that pre-existing cardiovascular disease and hypertension had a stronger effect on COVID-19 mortality than respiratory disease and obesity. While the differences could be due in part to fewer UKB subjects having comorbid COPD, obesity and diabetes, the observation is consistent with published papers. In a meta-analysis of seven studies including 1,576 COVID-19 positive patients, the most prevalent comorbidities were hypertension and diabetes, followed by cardiovascular disease and respiratory disease.[13] The findings for the association of older age and male sex with COVID-19 mortality are consistent with previously published studies.[4,5,13] Different from previous studies,[14,15] we did not find the association of the two genetic markers (ABO blood type and APOE) with COVID-19 positivity and mortality.

Several limitations of this study are noted. One is the relatively small sample size of tested subjects for COVID-19, which limited statistical power. A related limitation is that none of the reported P-values were corrected for multiple testing. Therefore, both positive and negative results of the study should be interpreted with caution. Additional released COVID-19 data from the UKB will be helpful to address both limitations. Another major limitation is the small number of minority subjects in the UKB. Finally, the lack of detailed clinical information on COVID-19 symptoms/treatment and cancer treatment limited our ability to perform a more comprehensive analysis.

In conclusion, in this first association study of cancer with COVID-19 risk and mortality from a population-based cohort, we showed that cancer patients in general have lower observed risk for COVID-19 infection but higher risk for COVID-19 death after infection, especially for hematologic cancers. The lower risk for COVID-19 infection, most likely due to extra caution in COVID-19 prevention and more testing among cancer patients, is encouraging and demonstrates the feasibility of intervention. We also showed that a subset of cancer patients is more likely to die of COVID-19 after the infection. These results, if confirmed, may provide guidance for COVID-19 prevention and treatment among cancer patients. Stronger personal protection should be made for cancer patients, and more intensive surveillance and/or treatment should be considered when cancer patients are infected with COVID-19.

## Data Availability

All datasets used in this study are accessible through the UKB. For more information about data availability, please contact the corresponding author, Jianfeng Xu, MD, Dr.PH, at.

## Additional Information

The UKB was approved by North West – Haydock Research Ethics Committee (REC reference: 16/NW/0274; IRAS project ID: 200778). UKB data was accessed through a Material Transfer Agreement under Application Reference Number: 50295. This study was performed in accordance with the Declaration of Helsinki. All UKB participants gave their informed consent before any data/samples were collected.

## Acknowledgements

We are grateful to the Ellrodt-Schweighauser, Chez and Melman families for establishing Endowed Chairs of Cancer Genomic Research and Personalized Prostate Cancer Care at NorthShore University HealthSystem in support of Dr. Xu and Dr. Helfand, the Rob Brooks Fund for Personalized Prostate Cancer Care at NorthShore University HealthSystem.

## Author Contributions

Concept: Xu

Data analysis: Shi, Wei, Na, Wang

Manuscript draft: Shi, Resurreccion, Xu

Manuscript revision: All authors

